# Exploring Midwives’ Practice Patterns and Capacity for Obstetric Ultrasound Imaging: Towards a Multicentre Longitudinal Materno-foetal Research Readiness in a Low-Resource Setting

**DOI:** 10.1101/2025.08.13.25332486

**Authors:** Albert Dayor Piersson, Philomena Ajanaba Asakeboba, Sarah Teiko Quartei, Rachel Mendy, Ama Boahene Akomah, Gilbertson Allorsey

## Abstract

**Introduction:** Midwives are often the first point of contact for pregnant women; yet their roles, training, and referral practices regarding obstetric ultrasound vary widely. This study aimed to explore midwives’ perspectives and experiences with obstetric ultrasound across key clinical and operational domains to assess the feasibility of conducting future multicentre maternal-foetal health research and surveillance.

**Methods:** A descriptive cross-sectional study was conducted among 473 practicing midwives across diverse healthcare settings in Ghana. A self-administered structured questionnaire was used to collect data on midwives’ perspectives and experiences regarding obstetric ultrasound across multiple dimensions. Data analysis was performed using Microsoft Excel.

**Results:** Most midwives were female, aged 26–35 years, held diploma qualifications, and practiced within district hospitals. Key ultrasound measures prioritised by midwives in the 1^st^ trimester include gestational age, foetal viability, estimated date of delivery (EDD), number of fetuses, and the presence of an intrauterine gestational sac. Comparatively, midwives emphasize foetal anomaly detection, amniotic fluid (liquor) volume, placental location, foetal viability, and gestational age during second trimester ultrasound screening, while in the 3^rd^ trimester screening, they prioritise foetal presentation, amniotic fluid volume, estimated foetal weight, placental location, and foetal viability. Findings suggest infrequent ultrasound reports indicating foetal anomalies. We observed a moderate perceived ability among midwives to understand foetal anomalies on obstetric ultrasound reports. Only 57.5% indicated they refer patients between one and three times for obstetric ultrasound before delivery. From the findings, it was observed that there is a predominance of sonographers undertaking obstetric ultrasound scans. Midwives may have moderate competence in interpreting obstetric ultrasound reports. An overwhelmingly positive response indicated that obstetric ultrasound improved their work performance, and a high proportion expressed interest in learning how to undertake obstetric ultrasound.

**Conclusion:** Our findings highlight the need to standardize midwifery practices and strengthen obstetric ultrasound literacy through targeted capacity-building initiatives, not only to improve clinical decision-making but also to establish a robust foundation for scalable maternal-foetal research in low-resource settings. Additionally, our study demonstrates the potential feasibility of engaging midwives as key stakeholders in multicentre maternal-foetal research initiatives.

## Introduction

Obstetric ultrasound has become an indispensable component of antenatal care (ANC), offering critical insights into foetal development, maternal health, and pregnancy outcomes. In many healthcare settings, midwives are frontline providers of maternity services and play a crucial role in the coordination, interpretation, application of obstetric ultrasound findings, interventions and provision of appropriate treatments throughout pregnancy [1]. Their understanding of clinical indications for referral, competency in the interpretation of ultrasound reports, and recognition of foetal anomalies are vital for timely and appropriate care [2, 3].

The clinical indications for obstetric ultrasound are diverse and span the entirety of prenatal care. In the first trimester, ultrasound is commonly used for confirming viability, determining gestational age, and identifying multiple pregnancies [4]. In the second trimester, it is instrumental in anatomical surveys and the detection of congenital anomalies [5], while third-trimester scans are typically employed for growth monitoring and assessment of foetal wellbeing [6]. However, the extent to which midwives understand and apply these indications in practice, especially in resource-limited contexts, remains understudied.

As providers who often initiate referrals, midwives’ awareness of foetal anomalies and their frequency of observation in clinical practice can inform the urgency and justification for scans [3, 7]. Moreover, patterns in how frequently patients are referred for ultrasound prior to delivery may reflect both institutional protocols and individual clinical judgment. Competence in interpreting ultrasound reports is another critical area, particularly given that many midwives rely on second-hand interpretation to inform their clinical decisions. This raises questions about the impact of ultrasound on their work performance [8] and their confidence in using ultrasound findings to guide care [9].

This study investigates midwives’ perspectives and experiences regarding obstetric ultrasound across multiple dimensions—clinical indications, key trimester-specific ultrasound measures, detection of foetal anomalies, frequency of referrals, interpretation competence, perceived work impact, and their willingness to learn ultrasound skills. The findings aim to inform not only policymaking, education, and practice, but also to support the design and feasibility of future multicentre materno-foetal health research and surveillance.

## Methods

### Study Design and Setting

A cross-sectional descriptive study was conducted among practicing midwives across the regions in Ghana from 17^th^ January – 28^th^ April, 2025. The study aimed to explore referral patterns, clinical decision-making, and knowledge of obstetric ultrasound use, with an additional focus on understanding midwives’ awareness of foetal anomalies, interpretation of ultrasound reports, and their willingness to be trained in basic obstetric ultrasound. The study was designed to generate context-specific data that could inform maternal health policy and lay the foundation for multicentre materno-foetal research in Ghana.

### Participants and Sampling

A total of 473 registered midwives working in primary, secondary, and tertiary healthcare facilities were recruited. Purposive sampling was employed to ensure a diverse representation of midwives from urban, peri-urban, and rural regions, including district hospitals and regional health centres. Eligibility criteria included being actively involved in ANC and willing to provide informed consent.

### Data Collection Instrument

Data were collected using a structured, self-administered questionnaire developed by the research team based on a literature review and input from maternal health experts. The questionnaire covered the following: demographic characteristics (age, gender, years of practice, qualification, facility type, and region), clinical indications considered important for obstetric ultrasound referrals, key ultrasound parameters reviewed during first, second, and third trimester screening, frequency of patient referrals for ultrasound prior to delivery, frequency of foetal anomalies observed in practice, roles typically involved in conducting obstetric ultrasound, knowledge and interpretation of ultrasound reports, perceived impact of ultrasound on midwifery work performance, and willingness to undertake training in obstetric ultrasound. The tool was piloted among 20 midwives in a non-study region to assess clarity and relevance. Minor modifications were made based on feedback, ensuring face and content validity.

**Table 1.** Interpretation of knowledge of interpretation of obstetrics ultrasound report, understanding of foetal anomalies on obstetric ultrasound report, and referral frequencies.

| knowledge of interpretation of obstetrics ultrasound report and understanding of foetal anomalies on obstetric ultrasound report |  | Referral Frequencies |  |
| --- | --- | --- | --- |
| Score Range | Interpretation | Referral Frequency Group | Range |
| Low knowledge | 1 - 3 | Low | 1-3 times |
| Moderate knowledge | 4 - 6 | Moderate | 4-6 times |
| High knowledge | 7 - 10 | High | 7-10 times |

### Data Analysis

Data were entered into a Google form and exported to Microsoft Excel for statistical analysis. Descriptive statistics (frequencies, percentages, means, and standard deviations) were used to summarise variables. Knowledge of interpretation of obstetrics ultrasound report and understanding of foetal anomalies on obstetric ultrasound report were grouped into three score categories: 1–3, 4–6, and 7–10; while referral frequencies were grouped into three categories: 1–3 times, 4–6 times, and 7– 10 times (**Table 1**). The mean, mode, and range of referral frequencies were calculated to describe common patterns of ultrasound use. For items with multiple-response options (e.g., ultrasound roles or clinical indications), response frequencies were calculated. Frequency distributions were visualised using pie and bar charts in Excel to illustrate distribution patterns and key findings.

### Ethical Considerations

Ethical approval for this study was obtained from the Centre for Education, Population Health Research & Innovation (ID: CEPHRI/ERC/2025/0781). Midwife WhatsApp group administrators across various regions were contacted to disseminate a self-administered questionnaire, designed using Google Forms, via their platforms. To ensure broader participation—especially in areas with limited internet access—hard copies of the questionnaire were also distributed. Additionally, faculty members who engaged with midwives during seminars and workshops facilitated further dissemination across regions. Participation was entirely voluntary, and written informed consent was obtained from all respondents. To maintain confidentiality, data were anonymised and used exclusively for research purposes.

## Results

**Table 2.** Demographics.

| Items | Midwives, n = 473 |
| --- | --- |
| Age (years) |  |
| 18-25 | 67 (14.4) |
| 26-30 | 145 (31.3) |
| 31-35 | 135 (29.1) |
| 36-40 | 89 (19.2) |
| ≥ 41 | 30 (6.5) |
| No response | 9 |
| Sex |  |
| Male | 19 (4.1) |
| Female | 440 (95.9) |
| No response | 14 |
| No of years in midwifery practice |  |
| 0-5 | 275 (59.1) |
| 6-10 | 159 (34.2) |
| 11-15 | 22 (4.7) |
| 16-20 | 7 (1.5) |
| ≥ 21 | 2 (0.4) |
| No response | 7 |
| Current midwifery qualification |  |
| Certificate | 45 (9.7) |
| Diploma | 351 (76.0) |
| BSc | 63 (13.6) |
| Postgraduate | 2 (0.4) |
| Other | 4 (0.9) |
| No response | 11 |
| Type of facility of work |  |
| CHPS | 29 (6.3) |
| Health centre | 82 (17.9) |
| Municipal Hospital | 85 (18.5) |
| District Hospital | 160 (34.9) |
| Regional Hospital | 22 (4.8) |
| Teaching Hospital | 40 (8.7) |
| Private Hospital | 34 (7.4) |
| CHAG | 4 (0.9) |
| Others | 5 (1.1) |
| No response | 14 |
| Region of work location |  |
| Ashanti | 8 (1.7) |
| Central | 81 (17.6) |
| Savanna | 16 (3.5) |
| Volta | 6 (1.3) |
| Bono | 11 (2.4) |
| Eastern | 8 (1.7) |
| North East | 14 (3.1) |
| Oti | 6 (1.3) |
| Bono East | 8 (1.7) |
| Greater Accra | 79 (17.2) |
| Upper East | 30 (6.5) |
| Western | 0 (0) |
| Ahafo | 2 (0.4) |
| Northern | 165 (35.9) |
| Upper West | 25 (5.4) |
| Western North | 2 (0.7) |
| No response | 14 |

The demographic profile (Table 2) of the study participants revealed that the majority of midwives were within the 26–35 years age range, with over 95% identifying as female. Most respondents had between 0–5 years of clinical experience, held a Diploma qualification in midwifery, and were predominantly employed in district-level hospitals. A significant proportion of the participants were based in the Northern region, reflecting the geographical distribution of midwifery practice within the study context.

**Figure 1.**
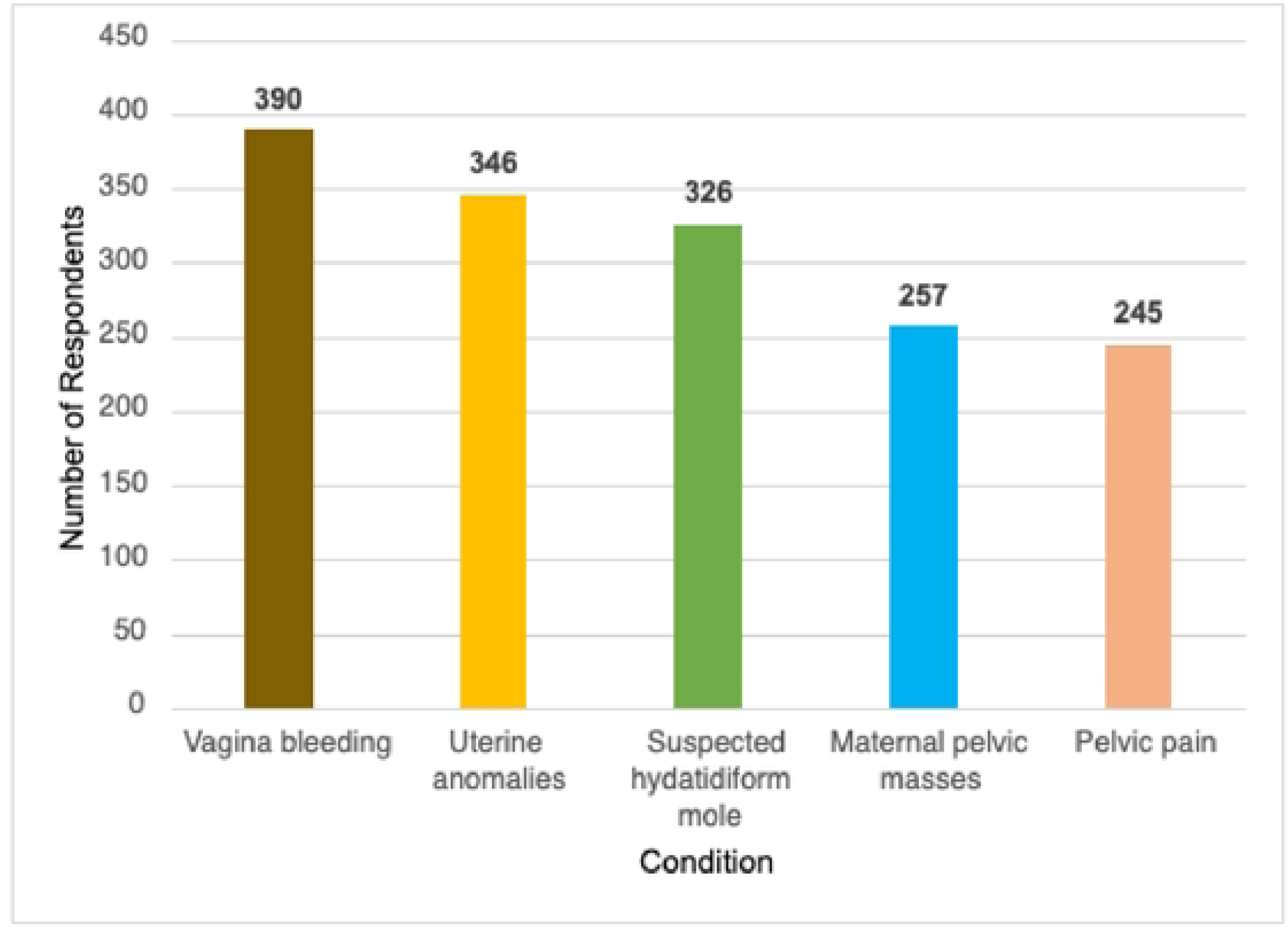
Clinical indications considered important for obstetric ultrasound

**Figure 2.**
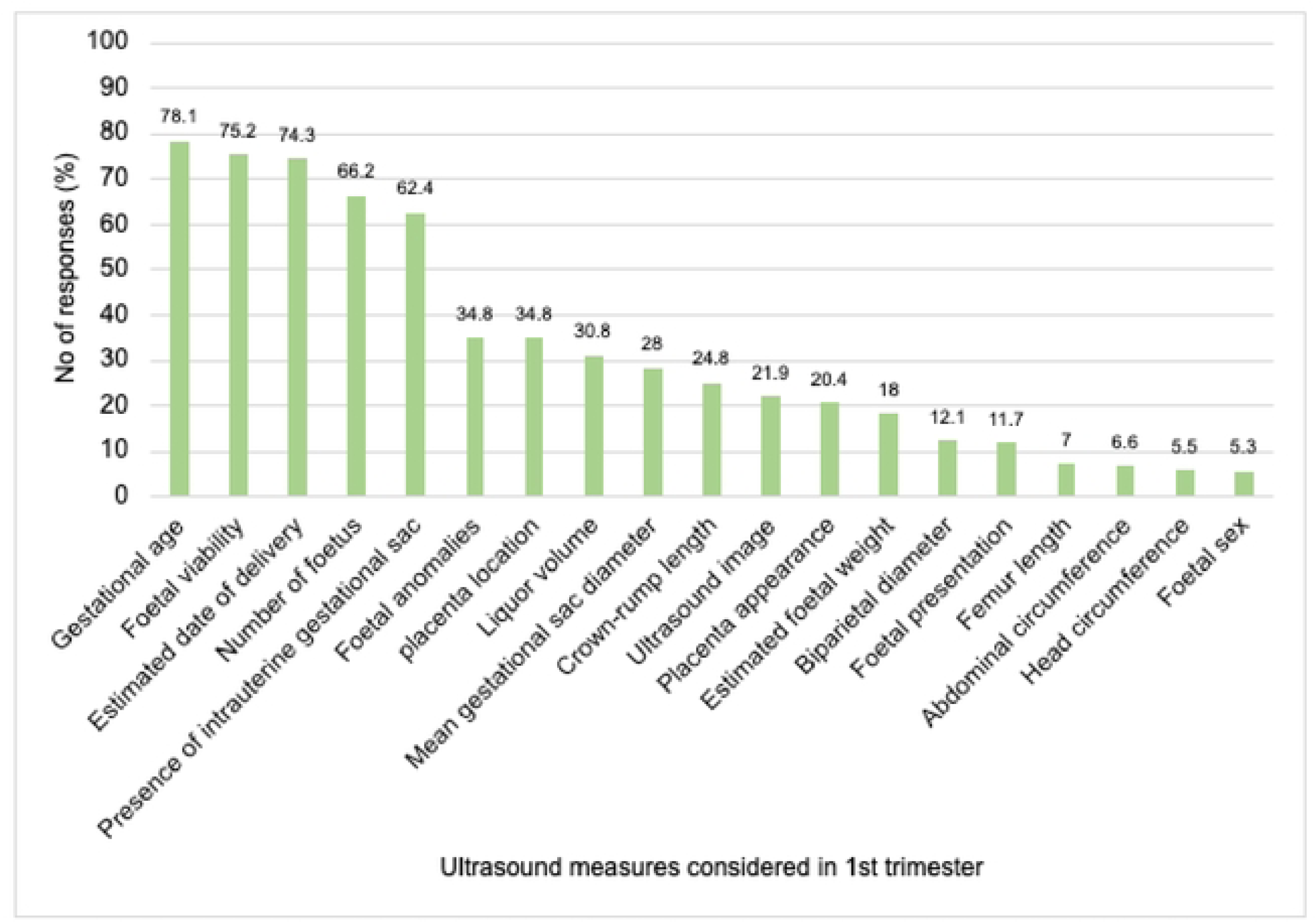
Key Ultrasound Measures Considered by Midwives During 1st Trimester Prenatal Screening

**Figure 3.**
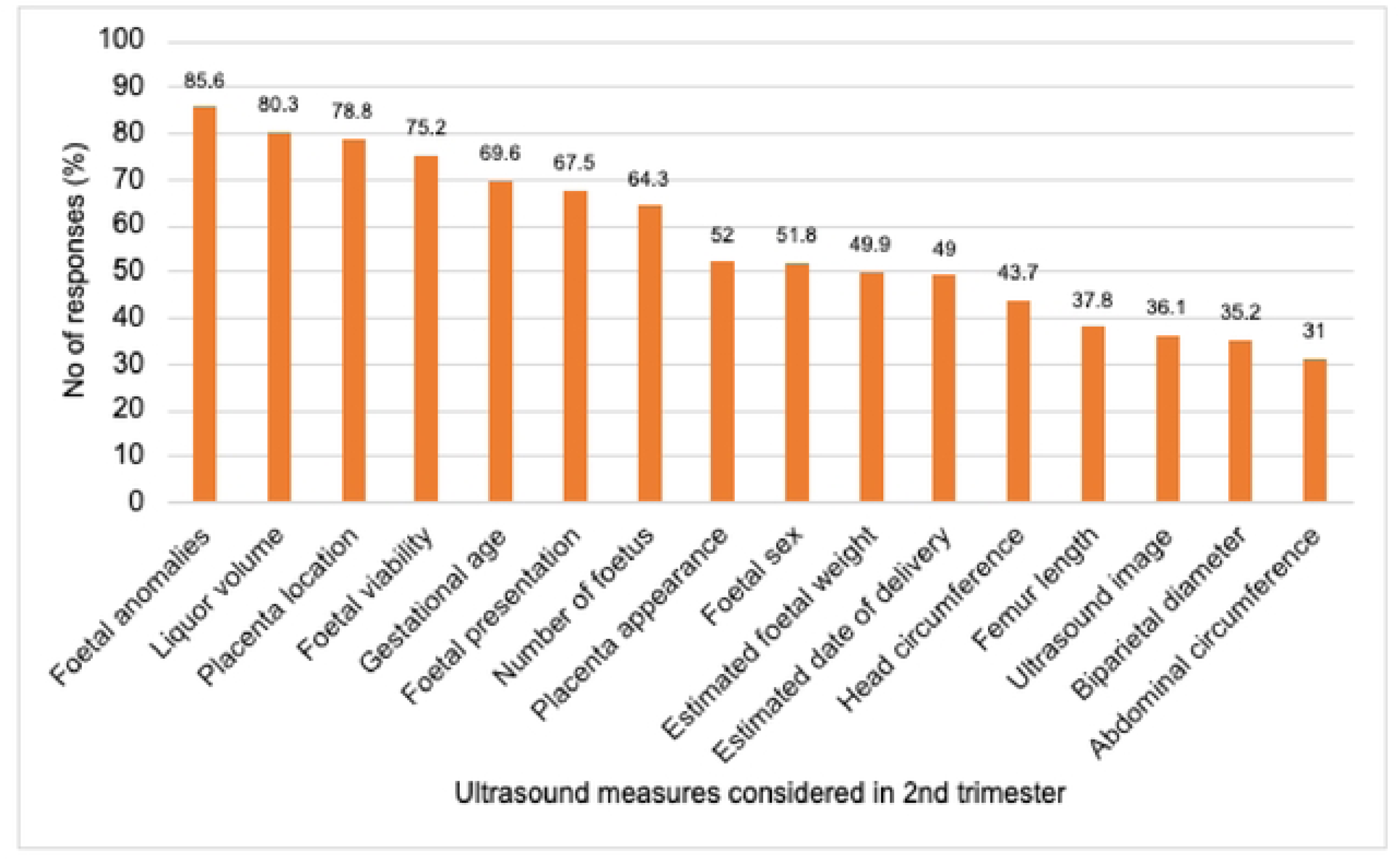
Key Ultrasound Measures Considered by Midwives During 2nd Prenatal Screening

**Figure 4.**
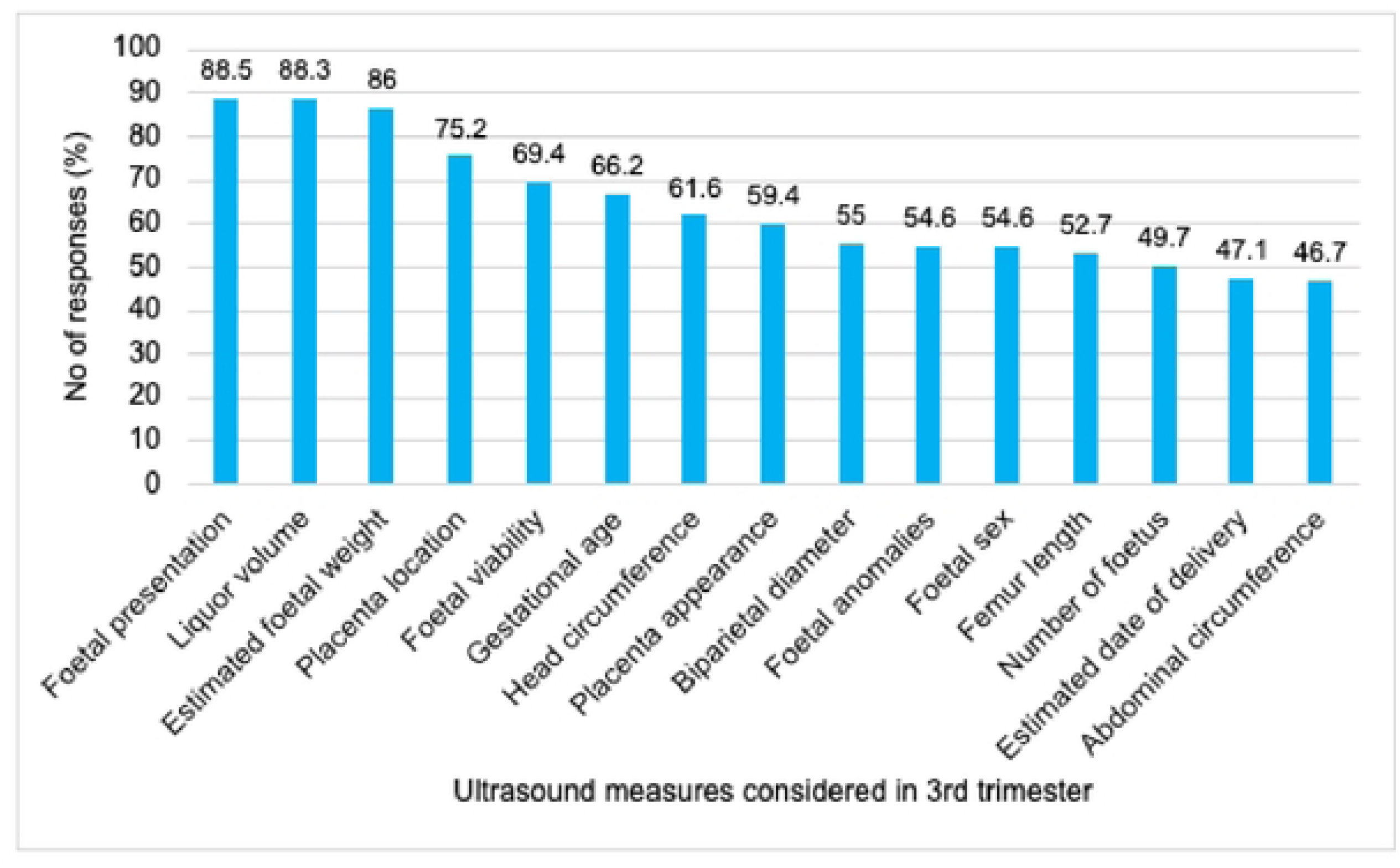
Key Ultrasound Measures Considered by Midwives During 3rdTrimester Prenatal Screening

**Figure 5.**
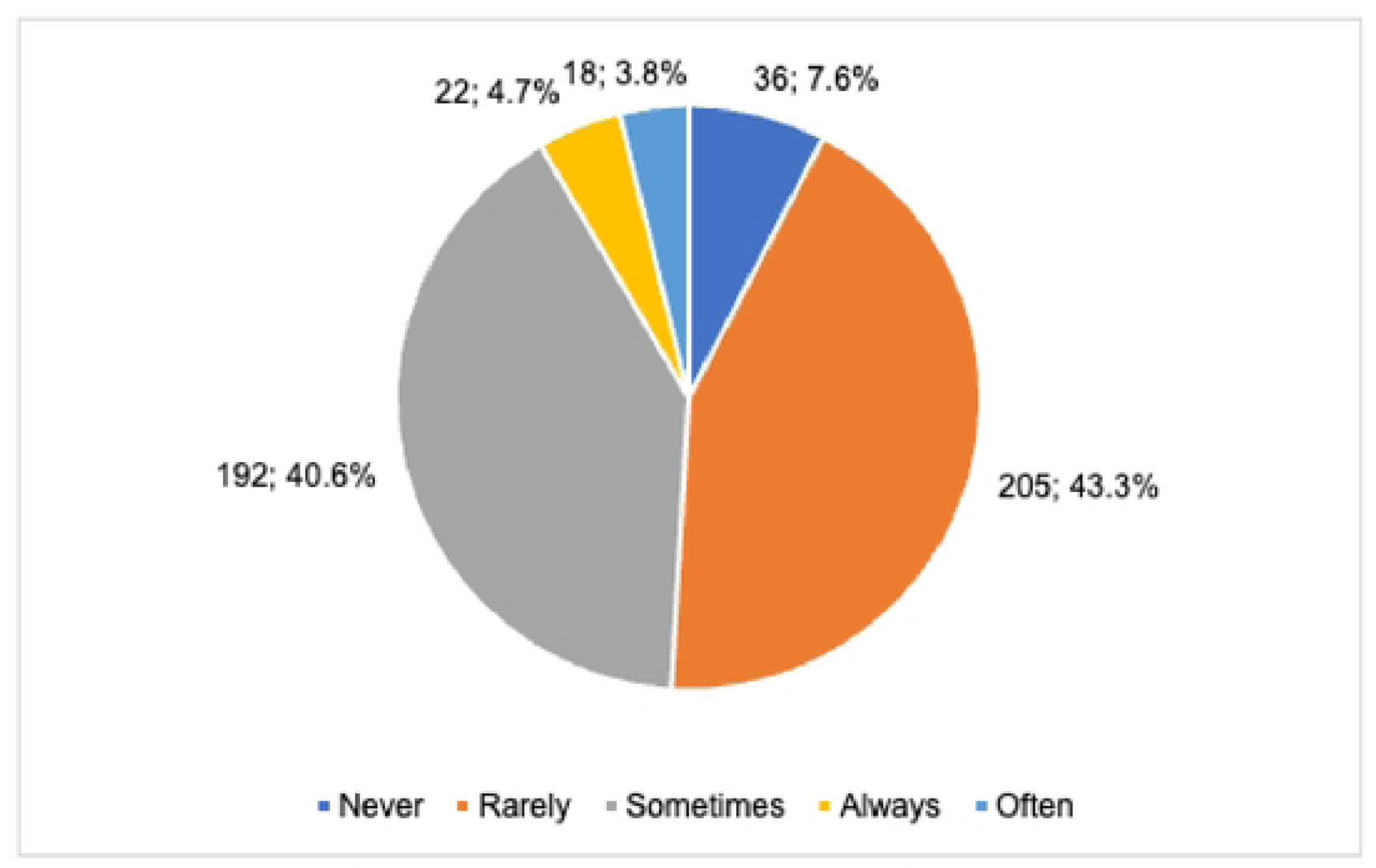
Frequency of foetal anomalies observed in practice

**Figure 6.**
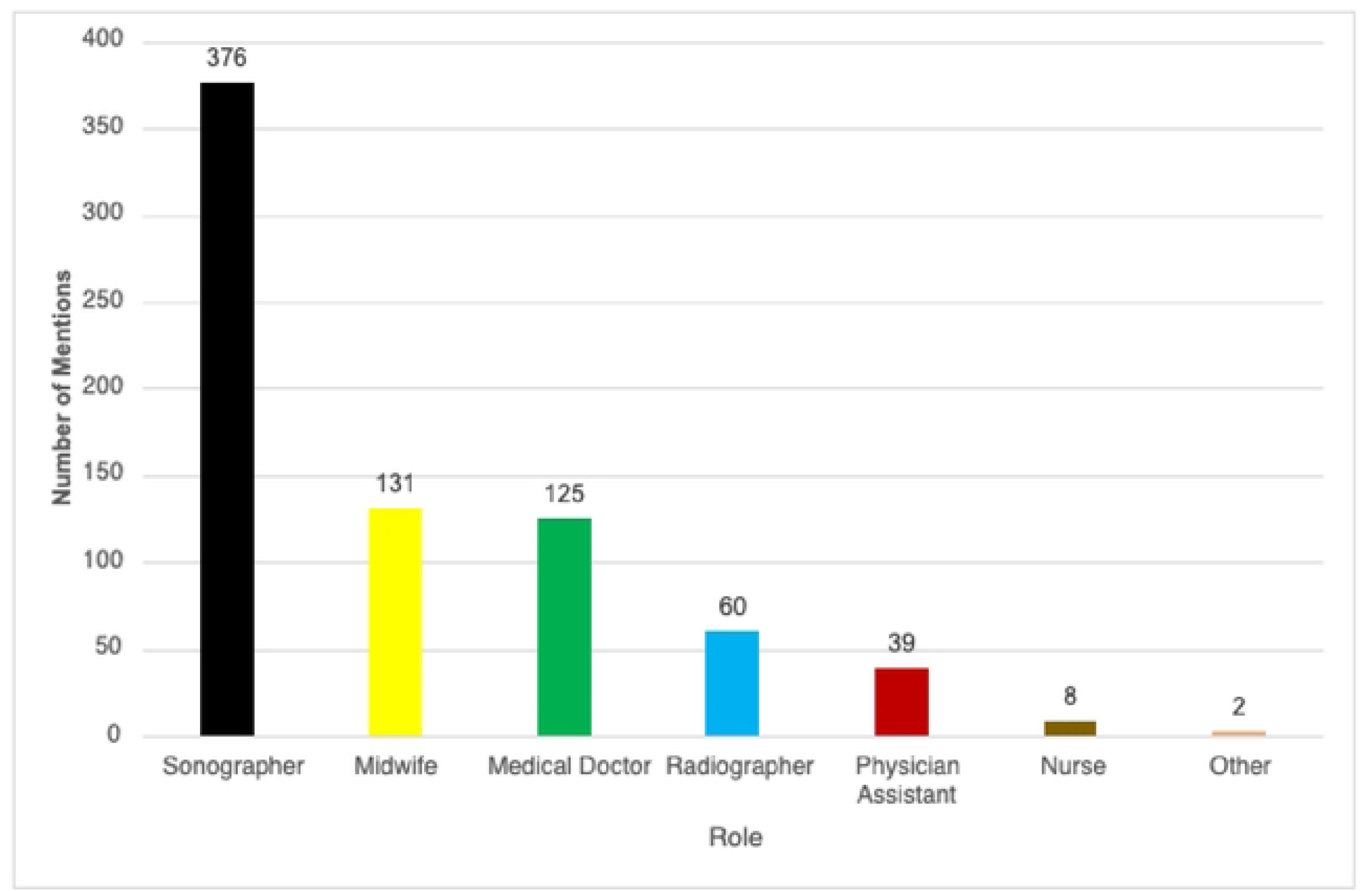
Frequency of roles performing obstetric ultrasound

**Table 3.** Knowledge of obstetric ultrasound report interpretation, awareness of foetal anomalies, and frequency of patient referrals prior to delivery.

| Score Range | Knowledge of interpretation of obstetrics ultrasound report |  | Understanding of foetal anomalies on obstetric ultrasound report |  | Frequency of patient referrals for obstetric ultrasound prior to delivery |  |  |
| --- | --- | --- | --- | --- | --- | --- | --- |
|  | n (%) | Mean ± SD | n (%) | Mean ± SD | n (%) | Weighted Average | Mode |
| 1 – 3 | 111 (23.5) | 5.19 ± 2.25 | 127 (26.9) | 5.11 ± 2.26 | 272 (57.1) | 4.44 | 3 |
| 4 – 6 | 219 (46.3) |  | 253 (53.5) |  | 106 (22.4) |  |  |
| 7 – 10 | 143 (30.2%) |  | 93 (19.7) |  | 95 (20.1) |  |  |

**Figure 7.**
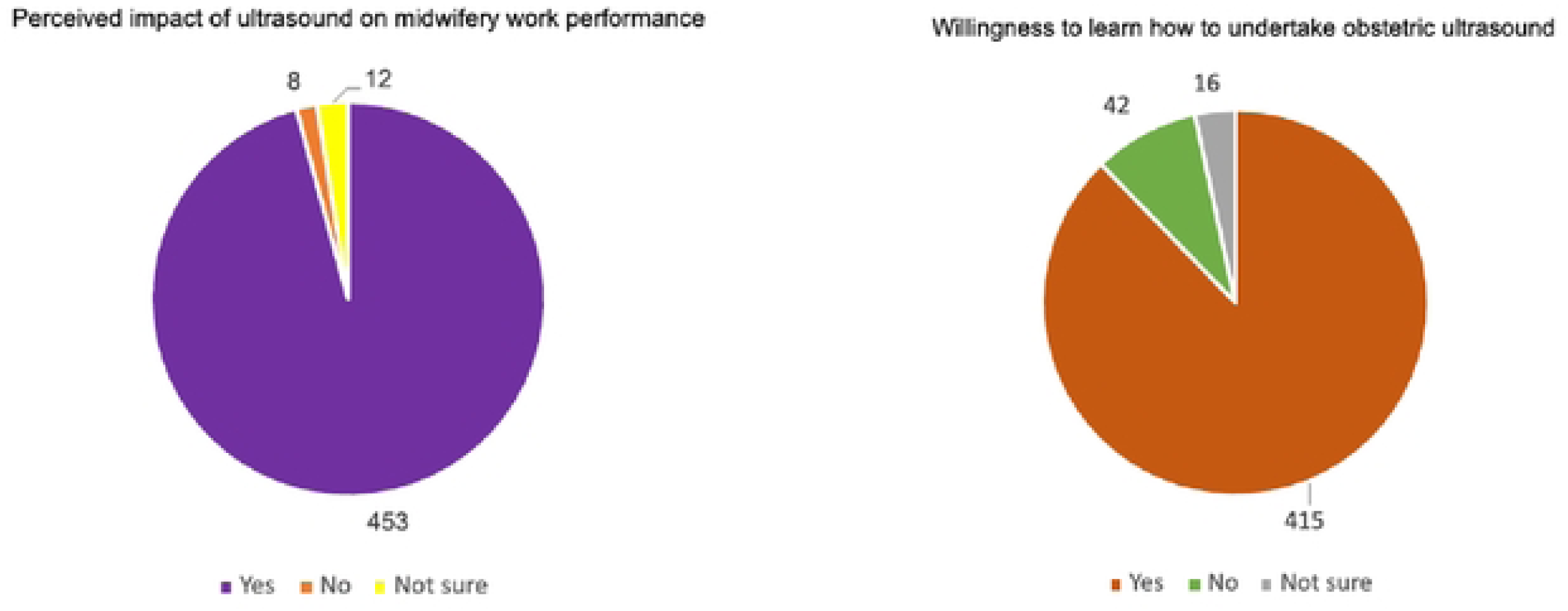
Perceived impact of ultrasound on midwifery work performance and willingness to learn how to undertake obstetric ultrasound.

## DISCUSSION

### Clinical Indications for Obstetric Ultrasound

When we asked midwives to pinpoint the indications they consider important for obstetric ultrasound besides routine antenatal scanning, they prioritized vagina bleeding, uterine anomalies, and suspected hydatidiform moles (**Figure 1**). This highlights a clinical focus on high-risk maternal conditions with potential for adverse pregnancy outcomes—an essential strategy in LMICs where late presentation and limited access to specialist care persist. The prevalence of medically complicated pregnancies is high among women living in LMICs [10], and this has been linked to late accessing of ANC [11]. The pattern demonstrated in our study reflects both clinical necessity and their awareness which clearly underscores the importance of ensuring timely and accurate sonographic evaluation to support maternal–foetal health surveillance.

### Key Ultrasound Measures Considered by Midwives

The study findings revealed that midwives prioritise gestational age, foetal viability, estimated date of delivery (EDD), number of fetuses, and the presence of an intrauterine gestational sac as key ultrasound measures, they look out for in the first trimester (**Figure 2**). This aligns with international recommendations on the essential components of early antenatal ultrasound screening [4, 12]. These parameters are critical for accurately dating pregnancies, identifying multiple gestations, and detecting early pregnancy failures or ectopic gestations—conditions that, if undiagnosed, contribute significantly to maternal and perinatal morbidity and mortality in LMICs. We found a low consideration for crown-rump length in the first trimester despite its relevance. Crown-rump length, which is optimally measured after 10 weeks’ gestation but before 14 weeks gestation can be used to estimate gestational age in the first trimester and it is considered superior to gestational sac diameter for gestational age estimation [13, 14]. Towards the end of the first trimester, obstetric ultrasound also offers an opportunity to detect major foetal abnormalities and, in healthcare systems that offer first-trimester aneuploidy screening, to measure the nuchal translucency (NT) thickness [4]. NT screening is now being offered routinely in Western countries [15, 16] as opposed to LMICs; however, it is feasible [17]. While we did not indicate NT in our study, the extent to which it is undertaken as part of routine first trimester scan in local settings is not well established and worths exploring in future studies. However, we speculate it is not routinely undertaken.

Comparatively, midwives place emphasis on foetal anomaly detection, amniotic fluid (liquor) volume, placental location, foetal viability, and gestational age during second trimester ultrasound screening (**Figure 3**), reflecting adherence to global best practices. The second trimester ultrasound provides a key opportunity for evaluating foetal development and identifying structural abnormalities [5, 18]. Interestingly, low priority was placed on the biparietal diameter (BPD), head circumference (HC), abdominal circumference (AC) and femur length (FL) which can be measured routinely for the assessment of foetal size. Perhaps this may be due to the lack or inadequate knowledge in understanding these measures, their contribution to antenatal assessment, and how to apply them in ANC. These four parameters provide information on the estimated foetal weight [5]. Besides, while gestational age is one of the key measures midwives prioritise in this trimester, they should also be aware that if a fetus has not been dated previously, HC or HC plus FL can serve as alternative measure(s) for dating after 14 weeks [5]. In LMICs, where access to tertiary diagnostic services may be limited, the accurate assessment of these parameters is crucial for early detection of complications such as oligohydramnios, placenta previa, and congenital anomalies, enabling timely referrals and improved pregnancy outcomes. However, this can only be achieved with optimized coordination among interprofessional team members [19]. As such, it is recommended that health professionals involved in second-trimester obstetric sonography must possess specific technical skills, which includes proficiency in operating ultrasound equipment, identifying foetal structures, and interpreting sonographic images accurately [19].

In the third trimester screening, midwives prioritise foetal presentation, amniotic fluid volume, estimated foetal weight, placental location, and foetal viability (**Figure 4**), reflecting appropriate clinical focus on parameters essential for intrapartum planning and the prevention of complications such as obstructed labour, foetal distress, and stillbirth. This aligns with international guidelines [6, 12], which emphasise the value of third trimester ultrasound in late pregnancy risk stratification, especially in LMICs where clinical resources are often limited and timely identification of high-risk pregnancies can significantly improve maternal-foetal outcomes. Besides these measures, the third-trimester ultrasound scan can also be used to assess fetoplacental Doppler and it is useful for assessment of gestational age if the woman has not had a previous ultrasound scan [6].

### Frequency of Foetal Anomalies Observed in Practice and Awareness of Foetal Anomalies

We also explored the frequency of midwives receiving obstetric ultrasound report indicating foetal anomalies (**Figure 5**). We observed the predominance of “rarely” and “sometimes” responses, suggesting that reports indicating foetal anomalies are relatively infrequent in routine obstetric ultrasound practice, which may reflect low detection rates, true low prevalence, or even under-reporting. But Swarray-Deen et al [20] highlighted the substantial burden of congenital anomalies detected through prenatal ultrasound at a tertiary referral center in Ghana. Frequent anomalies reported in Ghana are not limited to ventriculomegaly, anorectal malformations, cleft lip and cleft palate, spina bifida, exomphalos, and talipes [3, 20, 21]. Therefore, the trend revealed in our study may highlight a potential gap in early anomaly detection and standardized anomaly reporting. We also observed a moderate perceived ability among midwives to understand foetal anomalies on obstetric ultrasound reports (**Table 3**). This compares with a study in Ghana regarding midwives’ ability to identify neonates with congenital anomalies [3]. Cumulative evidence suggests that the presence of foetal anomalies is associated with adverse maternal health outcomes [22] including the risk of depression and traumatic stress in expectant parents, both acutely and over time [23]. A study that assessed the general public’s knowledge of fatal foetal anomalies identified the lack of accurate knowledge on them, their classification, diagnosis, and survival [24]. More so, women’s knowledge on congenital anomalies is reported to be limited [25, 26]. In LMICs, where access to specialised foetal medicine training and continuous professional development is often limited, gap in knowledge could lead to under-recognition or misinterpretation of congenital anomalies, potentially affecting early intervention and referral pathways. Therefore, capacity building in recognising and understanding foetal anomalies is necessary to enhance the feasibility and data quality of future multicentre maternal–foetal research.

### Frequency of Patient Referrals Prior to Delivery

A critical analysis of the findings reveals that a growing reliance on obstetric ultrasound is evident among midwives, with 57.5% referring patients between one to three times prior to delivery (**Table 3**). This trend surpasses the World Health Organization’s minimum recommendation of one ultrasound before 24 weeks’ gestation, reflecting an increasing integration of ultrasound into routine ANC. The study reported a mean referral frequency of four, which aligns with previous findings by Zile-Velika et al [27], who observed an average of 3–4 scans per pregnancy. However, this apparent uptake is marked by notable variation in referral patterns across midwives, pointing to inconsistencies in clinical practice. Several underlying factors may contribute to this heterogeneity, including differences in clinical experience, variable access to ultrasound services, under-resourced facilities, and institution-specific protocols that guide referral behaviours. Additionally, late presentation for ANC and divergent levels of midwives’ confidence in interpreting and utilising ultrasound findings can affect decision-making around the timing and frequency of referrals. Patients also play a critical role in shaping referral patterns for obstetric ultrasound and cannot be entirely absolved from the observed variability. Several interconnected factors can influence midwives’ decisions, including patients’ geographical access and proximity to ultrasound facilities, socioeconomic constraints, cultural and personal beliefs, individual preferences or anxieties, the influence of partners or families, late presentation to ANC. Other studies have also highlighted some of these factors [28, 29]. The referral pattern demonstrated in our study somehow reflects improved access and the perceived clinical value of ultrasound in monitoring maternal and foetal health. This growing uptake may be partly attributed to Ghana’s Free Maternal Healthcare Policy, introduced under the National Health Insurance Scheme in 2008, which aimed to reduce financial barriers to essential maternal services. However, its implementation has also been criticizedd for not fully eliminating out-of-pocket costs [30–32]. But we also observed another varied distribution, with nearly 43% of midwives referring patients four or more times for obstetric ultrasound. We consider that this may be due to pregnant women’s increasing demand for imaging, cases of assaults or trauma, post-term pregnancies, monitoring of high-risk pregnancies, fragmented continuity of care, uncertainty in gestational dating, low confidence in initial scan results, training and protocol gaps, financial incentives in private practice, inadequate regulation of ultrasound. This has implications for health policy. As such, these patterns underscore the importance of considering budget allocation for a number of potential research participants (∼ 40%) who may require more frequent obstetric ultrasound imaging (≥ 4 times). Given evidence that perinatal deaths increase with only four ANC visits and that an increase in the number of ANC contacts is associated with an increase in maternal satisfaction, consideration for this specific budget cover would align with the WHO’s [33] recommendation of a minimum of eight ANC contacts—one in the first trimester, two in the second, and five in the third.

### Frequency of Roles Performing Obstetric Ultrasound

We observed the predominance of sonographers undertaking obstetric ultrasound scans (**Figure 6**) reflecting government’s concerted effort to train to mitigate critical workforce shortages and ensure timely access to imaging services. In Ghana, for more than 5 years now, a few tertiary institutions have been running undergraduate and postgraduate Diagnostic Sonography programme, which may have contributed to the pattern observed in our study. However, for a potential multicentre maternal–foetal research, it is imperative to integrate highly specialized sonographers with a unique and advanced skill set comprising technical expertise in undertaking neurosonography and fetal echocardiography [34, 35]. Besides, there are documented positive impact of the provision of brief ultrasound training courses on local practices, particularly when provided to physicians [36, 37]and midwives [3]. But concerns have also been raised regarding the scale and proliferation of unlicensed sonographers and the impact on Ghana’s health care delivery [38]. While ultrasound is an intervention that can potentially be task shifted from trained sonographers to trained midwives, nurses, and other health professionals [12], we are also being critical of the depth of such short training, skill, knowledge and staff retention, and the extent to which follow-up refresher trainings are provided, and the overall impact on local practice. In addition, we are also concerned about the potential for interprofessional variability in scanning quality and report interpretation. These are important considerations for protocol standardisation and training in future multicentre maternal–foetal research initiatives in LMICs.

### Knowledge of Obstetric Ultrasound Report Interpretation

Our findings also indicate midwives may have a moderate competence in interpreting obstetric ultrasound reports (**Table 3**), highlighting potential variability in training and confidence levels across clinical settings. This compares with studies in other African countries that have reported a high performance in carrying out obstetrics ultrasound [39, 40], suggesting a corresponding high knowledge of the interpretation. However, the moderate knowledge evidenced from our study may reflect the lack or limited structured ultrasound education in midwifery curricula, posing a barrier to optimal utilization of ultrasound findings in clinical decision-making. There is the indication that the training of midwives on obstetric ultrasound scans in some African countries remains a serious challenge; however, there is a need to incorporate obstetric ultrasound scans as part of the scope of practice of midwives [1]. For instance, opportunities abound to adopt training curricula on basic scans that is inclusive of didactic and supervised clinical practicum components [41]. Addressing these knowledge gaps is critical not only for improving clinical outcomes but also for ensuring the reliability of data collection in prospective multicentre maternal–foetal research initiatives in LMICs.

### Perceived Impact of Ultrasound on Midwifery Work Performance and Willingness to Learn How to Undertake Obstetric Ultrasound

We also observed an overwhelmingly positive response indicating that obstetric ultrasound improved their work performance in midwifery practice (**Figure 7**), underscoring its pivotal role in enhancing clinical decision-making, timely referral, and overall maternal-foetal care. This finding aligns with studies in other African countries [8, 42] suggesting that access to diagnostic ultrasound strengthens midwives’ confidence in antenatal assessments, supports accurate gestational dating, and facilitates early detection of complications. However, such impact is often mediated by the quality of training, availability of equipment, and integration into standard protocols, emphasizing the need for continued professional development and infrastructural support in resource-limited settings. We also observed that a high proportion of respondents expressed interest in learning how to undertake obstetric ultrasound (**Figure 7**), demonstrating a strong motivation among frontline maternal healthcare providers to enhance their diagnostic capabilities, potentially driven by perceived clinical benefits and existing skill gaps. However, in a study, Holmlund et al [2] reported that midwives expressed that obstetric ultrasound scan was the duty of a physician and expressed discomfort in performing obstetric ultrasound citing safety concerns for the foetus. In another study, Argaw et al. [42] also raised concerns about increased workload, resulting from task shifting and extension of the scope of practice for midwives. As previously indicated, the establishment of undergraduate Sonography programmes in tertiary institutions may have helped improved the shortage of workforce in performing ultrasound. However, in Ghana, it is well-acknowledged that some medical professionals have refused postings to rural areas due to concerns about the conditions of service, perceived unfairness in the system, lack of opportunities, and lack of transparency in the posting process [43, 44]. But to enhance the representativeness and inclusivity of maternal–foetal research in LMICs, it is imperative to integrate underrepresented populations, particularly those in remote or underserved settings. Therefore, in line with the WHO’s recommendation [33], in the main time, while the sonography education steadily continues to grow the workforce, strengthening the capacity of midwives through targeted training in basic obstetric ultrasound presents a viable strategy to bridge this gap. However, there would be the need to complement such training initiatives with structured follow-up and periodic refresher programmes aimed at improving clinical accuracy and long-term competency, reinforcing skills, promoting quality assurance, and mitigating diagnostic variability.

### Strengths and Limitations

Our study is one of the few to systematically explore obstetric ultrasound practices from the perspective of midwives in LMIC settings, offering rich, context-specific insights relevant for maternal-foetal health system strengthening. Its large, multi-facility sample enhances the generalisability of findings across diverse clinical environments.

There are also limitations worth highlighting in our study. A key limitation of this study lies in its reliance on self-reported data, which may be subject to recall bias, social desirability bias, and variability in practice. In addition, we did not compare responses between facilities, and we did not independently verify clinical practice or diagnostic accuracy. Further, we focused solely on midwives. Finally, our study did not consider several other areas of midwifery practices.

### Conclusion and Implication for Health Policy and Prospective Multicentre Research Practices

Our study suggests the potential readiness and feasibility of engaging midwives as critical stakeholders in multicentre maternal-foetal research in a low-resource setting, where their clinical input and strategic use of ultrasound can enhance early risk detection and improve pregnancy outcomes. To inform health policy in LMICs, we recommend investment in structured, competency-based ultrasound training for midwives and other frontline providers, supported by national guidelines and quality assurance mechanisms. Future research can explore sonographers’ practices in relation to the performance of obstetrics ultrasound imaging. It would also be interesting to examine pregnant women’s views on care provided by midwives vis a vis their obstetric ultrasound report during ANC. Another area can be to assess the impact of communication strategies that promote shared decision-making during obstetric ultrasound appointments. Specifically, in line with the WHO [41], studies can examine how enabling pregnant women and their partners to view the ultrasound screen during discussions of foetal wellbeing may enhance understanding and engagement. Additionally, research should consider how health professionals acknowledge and manage women’s expectations and preferences regarding ultrasound within a respectful and informed care framework. Attention should be given to evaluating how well current practices convey the limitations of ultrasound, including its ability to predict foetal weight, mode of delivery, or detect certain anomalies—particularly in early pregnancy.

## Data Availability

All data produced in the present study are available upon reasonable request to the authors.

## References

1. Lukhele S, Mulaudzi FM, Sepeng N, Netshisaulu K, Ngunyulu RN, Musie M, et al. The training of midwives to perform obstetric ultrasound scan in Africa for task shifting and extension of scope of practice: a scoping review. BMC Medical Education. 2023;23(1).

2. Holmlund S, Ntaganira J, Edvardsson K, Lan PT, Sengoma JPS, Åhman A, et al. Improved maternity care if midwives learn to perform ultrasound: a qualitative study of Rwandan midwives’ experiences and views of obstetric ultrasound. Global Health Action. 2017;10(1).

3. Abdul-Mumin A, Rotkis LN, Gumanga S, Fay EE, Denno DM. Could ultrasound midwifery training increase antenatal detection of congenital anomalies in Ghana? PLoS ONE. 2022;17(8):e0272250.

4. ISUOG Practice Guidelines (updated): performance of 11–14-week ultrasound scan, Clinical Standards Committee, The International Society of Ultrasound in Obstetrics and Gynecology (ISUOG). ISUOG Practice Guidelines (updated): performance of 11–14-week ultrasound scan. Ultrasound Obstet Gynecol. 2023; 61:127–43.

5. Salomon LJ, Alfirevic Z, Berghella V, Bilardo CM, Chalouhi GE, Da Silva Costa F, et al. ISUOG Practice Guidelines (updated): performance of the routine mid-trimester fetal ultrasound scan. Ultrasound in Obstetrics and Gynecology. 2022;59(6):840–56.

6. ISUOG Practice Guidelines: performance of third-trimester obstetric ultrasound scan. Ultrasound Obstet Gynecol 2024; 63: 131–147

7. Gitsels-van der Wal JT, Manniën J, Gitsels LA, Reinders HS, Verhoeven PS, Ghaly MM, et al. Prenatal screening for congenital anomalies: exploring midwives’ perceptions of counseling clients with religious backgrounds. BMC Pregnancy and Childbirth. 2014;14(1).

8. Åhman A, Edvardsson K, Kidanto HL, Ngarina M, Small R, Mogren I. ‘Without ultrasound you can’t reach the best decision’ – Midwives’ experiences and views of the role of ultrasound in maternity care in Dar Es Salaam, Tanzania. Sexual & Reproductive Healthcare. 2017;15:28–34.

9. Daemers DOA, Van Limbeek EBM, Wijnen H a. A, Nieuwenhuijze MJ, De Vries RG. Factors influencing the clinical decision-making of midwives: a qualitative study. BMC Pregnancy and Childbirth. 2017;17(1).

10. McDonald CR, Weckman AM, Wright JK, Conroy AL, Kain KC. Pregnant women in low-and Middle-Income countries require a special focus during the COVID-19 pandemic. Frontiers in Global Women S Health. 2020;1.

11. Oduro CA, Opoku DA, Osarfo J, Fuseini A, Attua AA, Owusu-Ansah E, et al. The burden and predictors of late antenatal booking in a rural setting in Ghana. Nursing Open. 2022;10(4):2182–91.

12. Guidelines Review Committee. WHO recommendations on antenatal care for a positive pregnancy experience. 2016.

13. Pexsters A, Daemen A, Bottomley C, Van Schoubroeck D, De Catte L, De Moor B, et al. New crown–rump length curve based on over 3500 pregnancies. Ultrasound in Obstetrics and Gynecology. 2010;35(6):650–5.

14. WHO Recommendations on Antenatal Care for a Positive Pregnancy Experience: Ultrasound Examination. 2018. https://iris.who.int/bitstream/handle/10665/259946/WHO-RHR-18.01-eng.pdf

15. Bellai-Dussault K, Dougan SD, Fell DB, Little J, Meng L, Okun N, et al. Ultrasonographic fetal nuchal translucency measurements and cytogenetic outcomes. JAMA Network Open. 2024;7(3):e243689.

16. 16. Kozuma S. The dilemma surrounding nuchal translucency-thickness measurement in Japan J Med Ultrasonics. 2005 32:89–90

17. Oloyede OA, Abbey M, Oloyede AA, Nwachukwu O. Fetal nuchal translucency scan in Nigeria. Pan African Medical Journal. 2014 Jan 1;18.

18. Edwards L, Hui L. First and second trimester screening for foetal structural anomalies. Semin Foetal Neonatal Med 2018; 23: 102–111.

19. Jabaz D, Jenkins SM. Sonography 2nd Trimester Assessment, Protocols, and Interpretation. 2023

20. Swarray-Deen A, Yapundich M, Boudova S, Doffour-Dapaah K, Osei-Agyapong J, Sepenu P, et al. Spectrum of congenital anomalies detected through anatomy ultrasound at a referral hospital in Ghana. BMC Pregnancy and Childbirth. 2025;25(1).

21 Nii-Amon-Kotei D, Baffoe-Bonnie B. Congenital malformations in Kumasi, Ghana: prognosis for surgical cases. Pediatric Surgery International. 1991;6(1).

22. Atwani R, Aziz M, Saade G, Reddy U, Kawakita T. Maternal Implications of Fetal Anomalies: a Population-Based Cross-Sectional study. American Journal of Obstetrics & Gynecology MFM. 2024;6(10):101440.

23. Oftedal A, Bekkhus M, Haugen GN, Czajkowski NO, Kaasen A. The impact of diagnosed fetal anomaly, diagnostic severity and prognostic ambiguity on parental depression and traumatic stress: a prospective longitudinal cohort study. Acta Obstetricia Et Gynecologica Scandinavica. 2022;101(11):1291–9.

24. Power S, Meaney S, O’Donoghue K. An assessment of the general public’s knowledge of fatal fetal anomalies. Prenatal Diagnosis. 2018;38(11):883–90.

25. Kar A, Dhamdhere D, Medhekar A. “Fruits of our past karma”: a qualitative study on knowledge and attitudes about congenital anomalies among women in Pune district, India. Journal of Community Genetics. 2023;14(4):429–38.

26. Ferede AA, Kassie BA, Mosu KT, Getahun WT, Taye BT, Desta M, et al. Pregnant women’s knowledge of birth defects and their associated factors among antenatal care attendees in referral hospitals of Amhara regional state, Ethiopia, in 2019. Frontiers in Global Women S Health. 2023;4.

27. Zile-Velika I, Ebela I, Folkmanis V, Rumba-Rozenfelde I. Prenatal ultrasound screening and congenital anomalies at birth by region: Pattern and distribution in Latvia. European Journal of Obstetrics & Gynecology and Reproductive Biology X. 2023;20:100242.

28. Gashaw A, Figa Z, Abebe Y, Demeke AD, Sime Y. Knowledge and utilization of obstetric ultrasound and associated factors among pregnant mother in Africa: a systematic review and meta-analysis. The Ultrasound Journal. 2025;17(1).

29. Edzie EKM, Dzefi-Tettey K, Gorleku PN, Ampofo JW, Piersson AD, Asemah AR, et al. Perception of Ghanaian primigravidas undergoing their first antenatal ultrasonography in Cape Coast. Radiology Research and Practice. 2020;2020:1–10.

30. Dalinjong PA, Wang AY, Homer CSE. The operations of the free maternal care policy and out of pocket payments during childbirth in rural Northern Ghana. Health Economics Review. 2017;7(1).

31. Kumbeni MT, Afaya A, Apanga PA. An assessment of out of pocket payments in public sector health facilities under the free maternal healthcare policy in Ghana. Health Economics Review. 2023;13(1).

32. Piersson AD, Gorleku PN. Assessment of availability, accessibility, and affordability of magnetic resonance imaging services in Ghana. Radiography. 2017;23(4):e75–9.

33. WHO Recommendations on Antenatal Care for a Positive Pregnancy Experience: Summary. 2018 https://iris.who.int/bitstream/handle/10665/259947/WHO-RHR-18.02-eng.pdf

34. Stone J, Bromley B, Norton ME, Feltovich H, Platt LD, Copel JA, et al. Developing an optimal maternal-fetal medicine ultrasound practice: A report and recommendations of the workshop of the Society for Maternal-Fetal Medicine. Pregnancy. 2025; 1(3).

35. ISUOG Practice Guidelines (updated): sonographic examination of the fetal central nervous system. Part 2: performance of targeted neurosonography. Ultrasound Obstet Gynecol 2021; 57: 661–671

36. Osei-Ampofo M, Tafoya MJ, Tafoya CA, Oteng RA, Ali H, Becker TK. Skill and knowledge retention after training in cardiopulmonary ultrasound in Ghana: an impact assessment of bedside ultrasound training in a resource-limited setting. Emergency Medicine Journal. 2018;35(11):704–7.

37. Pathak A, Limbani F, Awuku YA, Booth A, Joekes E. Physicians’ clinical experience and perspectives following a pilot, blended learning, point of care ultrasound course in Ghana-a mixed methods analysis. BMC Medical Education. 2024;24(1).

38. Gorleku PN, Setorglo J, Ofori I, Edzie EKM, Dzefi-Tettey K, Piersson AD, et al. Towards the scale and menace of unregulated sonography practice in Ghana. Journal of Global Health Reports. 2020;4.

39. Swanson JO, Kawooya MG, Swanson DL, Hippe DS, Dungu-Matovu P, Nathan R. The diagnostic impact of limited, screening obstetric ultrasound when performed by midwives in rural Uganda. Journal of Perinatology. 2014;34(7):508–12.

40. Vinayak S, Brownie S. Collaborative task-sharing to enhance the Point-Of-Care Ultrasound (POCUS) access among expectant women in Kenya: The role of midwife sonographers. Journal of Interprofessional Care. 2018;32(5):641–4.

41. IMAGING ULTRASOUND BEFORE 24 WEEKS OF PREGNANCY: 2022 update to the WHO antenatal care recommendations for a positive pregnancy experience. 2022. https://iris.who.int/bitstream/handle/10665/362037/9789240051461-eng.pdf?sequence=1

42. Argaw MD, Abawollo HS, Tsegaye ZT, Beshir IA, Damte HD, Mengesha BT, et al. Experiences of midwives on Vscan limited obstetric ultrasound use: a qualitative exploratory study. BMC Pregnancy and Childbirth. 2022;22(1).

43. Okyere E, Ward P, Marfoh K, Mwanri L. What do Health Workers say About Rural Practice? Global Qualitative Nursing Research. 2021;8:233339362110548.

44. Kwamie A, Asiamah M, Schaaf M, Agyepong IA. Postings and transfers in the Ghanaian health system: a study of health workforce governance. International Journal for Equity in Health. 2017;16(1).

